# Swiss-Austrian VEry Preterm Infant Transition Systematic Review (SAVE-T): Transition to home: improving outcomes of children and families - a systematic review protocol

**DOI:** 10.1101/2024.04.23.24306012

**Authors:** Vivienne Engel, Valentina Mauron Papadimitriou, Lea Bührer, Laura Ebner, Lena Keller, Martina Gosteli, Sandra Siegfried, Beatrix Latal, Ursula Kiechl-Kohlendorfer, Ulrike Held

## Abstract

**Background:** Medical progress has significantly improved the survival rates of very preterm-born infants in recent decades. Nevertheless, these infants are still at increased risk for long-term impairments as compared to term-born infants. This situation often leaves parents coping with anxiety and a sense of unpreparedness, as they navigate the transition to home. While the homecoming of a preterm born infant is long awaited and brings relief to families, it also marks the end of intensive monitoring and highly specialized professional care. Correspondingly, the World Health Organisation (WHO) advocates for additional support for parents of preterm infants following the transition to home. According to the WHO, preparation for this transition yields positive effects on crucial aspects such as nutrition, parent-child interaction, and parental well-being. Consequently, we aimed to provide an overview of existing transition to home interventions and their efficacy, by conducting a systematic review of the literature.

**Methods:** We will perform a systematic review of interventions aiming at improving the transition to home process for very preterm-born infants and their parents and will search the following databases: Cochrane, Medline, CINAHL, EMBASE, and PsycINFO. Our main aim is to provide an overview over the existing interventions, and their efficacy. Thus, we will not predefine specific outcomes, but we will describe, assess and summarize the outcomes of the included studies.

**Discussion:** This systematic literature review aims to provide a comprehensive overview of current interventions designed to improve the transition from hospital to home. Subsequently, it will show the individual effect of these interventions and pool effect sizes if possible. The most promising interventions will then be combined and used as the basis of a subsequent randomized controlled trial of a new transition to home intervention.

**Systematic review registration:** The systematic review protocol was registered with the International Prospective Register of Systematic Reviews (PROSPERO) on August 30, 2023. PROSPERO CRD42023455401

## Background

The Federal Statistical Office of Switzerland states that 0,8% of births in 2021 were born very premature, that is below 32 weeks of gestation (1). This rate is comparable to other countries in Europe (2). According to the World Health Organization (WHO) preterm-born infants are at risk for severe morbidity and death (3). Their risk of mortality is two to ten times higher compared to term-born infants (4). Consequently, the birth of such an infant and the following hospital stay leads to emotional distress of the parents (5).

Very preterm-born infants have a higher risk of adverse outcomes during their development compared to moderate and late preterm-born and also term-born infants. These adverse outcomes affect different organ systems and can result in respiratory problems, hearing and visual impairment, gastrointestinal morbidities, neurodevelopmental disabilities, and cognitive impairment (6). During the neonatal hospital care, infants are constantly monitored and observed by medical staff. During this time, many interventions aim at improving parent-infant interaction (e.g. Kangaroo care, creative music therapy, breastfeeding etc.) and family centered care is performed in many neonatal units. The transition from hospital to home can be a vulnerable time for parents and their preterm-born infant. While coming home is long awaited and can be a relief for the families, it is also associated with an interruption of continuous or regular monitoring and highly specialized professional care.

Accordingly, a study conducted by Aydon et al. showed that parents of preterm-born infants felt anxious and not prepared regarding the transition to home (7). Furthermore, parents of very preterm-born infants often experience substantial stress which may also lead to symptoms of posttraumatic stress disorder (8). Correspondingly, in the “recommendations for care of the preterm or low birth weight infant” the WHO states that parents of preterm-born infants should receive additional support. This support should continue during and after the transition to home. They also stated that transition to home preparations had positive effects on nutrition, parent-child interaction and parental well-being (4).

We conclude that the transition to home process is an integral part of the development of a very preterm-born infant. Consequently, various studies and reviews are performed regarding this subject. For example Bedwell et al. are planning on performing a systematic review regarding “Interventions to support parents, families and carers in caring for premature or low birth weight infants in the home”. In contrast to our review, they will not focus on interventions that target the transition from hospital to home and they will also include low birth weight infants. They will focus on studies published in the last 20 years, whereas we will include studies published from 1990 onwards (9). Aagaard et al. published a systematic review protocol focusing on the parental experiences regarding the transition to home after discharge from the Neonatal Intensive Care Unit (NICU). They included all infants that were hospitalized in a NICU and defined a narrower time frame (2000-2014) in their search strategy compared to our systematic review (10).

Due to the importance, complexity and heterogeneity of the subject we aim to provide an overview of existing transition to home interventions for very preterm-born infants by conducting a systematic literature review and to summarize the efficacy of these interventions for different outcomes.

### Objectives

We aim to assess the existing literature of interventions focusing on improving the transition to home process for very preterm-born infants and their parents. These interventions may occur before, during or after hospital discharge and involve a subsequent follow-up assessment. If possible, we will compare these results with standard of care or no structured transition. Because our main goal is to work out an overview, we will not predefine or pre-select possible outcomes.

The result of this systematic review will serve as the foundation for a subsequent randomized controlled trial of a new transition to home intervention.

## Methods/Design

The Preferred Reporting Items for Systematic review and Meta-Analysis Protocols (PRISMA-P) 2015 checklist was used as a basic framework for this protocol. The checklist is added as **Additional file 1**. The systematic review protocol was registered with the International Prospective Register of Systematic Reviews (PROSPERO) on August 30, 2023 (registration number: CRD42023455401).

### Eligibility criteria

All studies that report transition to home interventions for very preterm-born infants (born before 32 0/7 weeks of gestation) and their parents.

### Study design

Any randomized controlled trial, observational cohort study, prospective study, retrospective study, case series with more than ten cases, case-control-study and systematic review will be included. Studies on case series with less than ten cases, conference abstracts and preprints will be excluded.

### Year of publication

We will include studies published from 1990 onwards, as this marks the starting point of the OSIRIS trial, a pivotal event that significantly increased the likelihood of survival for very preterm infants (11).

### Language

There will be no restrictions regarding language. During the full text screening studies in foreign languages will be translated with the translator program DeepL (www.deepl.com). The data extraction and quality assessment of these studies will be done with the help of a fluent speaker of the respective language, and with a medical background.

### Participants

Studies that report transition to home interventions for very preterm-born infants (born before 32 0/7 weeks of gestation) and their parents will be included.

Studies on full-term infants and on very preterm-born infants who still are in the hospital will be excluded. Studies focusing on infants with major congenital anomalies will be excluded. Studies on infants with low birth weight and other studies with heterogeneous populations where no specific information on very preterm-born infants can be retrieved will be excluded.

### Types of interventions and exposures

Any description of a structured hospital to home transition intervention for very preterm-born infants and their parents. These interventions may include individualized care plans, breastfeeding interventions, parental network groups, e-health services, phone calls and video conferences between neonatal health care professionals and parents, music therapy, Kangaroo Mother Care and other forms of interventions.

### Outcomes

#### Primary outcomes

The main goal of this review is to identify existing interventions improving the transition from hospital to home process for very preterm-born infants and their parents. Hence, we will not focus on specific predefined outcomes, but we will describe, evaluate and, if possible, summarize the outcomes of the included studies for different interventions. Consequently, the search strategy will not include pre-defined outcomes.

Expected outcomes regarding very preterm-born infants are behavior, self-regulatory abilities, neurodevelopmental outcome, growth, nutrition and health status. Expected outcomes regarding parents of very preterm-born infants are parent-child interaction, parameters of parental well-being and parental stress.

We will include follow-up assessments of outcomes with a maximum duration of two years.

#### Secondary outcomes

Health economic evaluations of transition to home will be considered as secondary outcomes.

### Information sources

#### Electronic database search

We will search the following biomedical databases: Cochrane library, Medline (EBSCOhost), CINAHL (EBSCOhost), EMBASE (embase.com), and PsycINFO (EBSCOhost).

We developed a search strategy for the following concepts: preterm AND transition to home AND parental support/need/interventions. For each concept we will use a combination of subject heading terms and free text words. We will not apply any restriction to language or study types. Benchmark studies will not be used. To minimize the risk of bias we will not use any search terms related to possible outcomes.

We are not planning on re-running the search prior to the final analysis. Unpublished studies will not be sought.

#### Search strategy

The search strategy was developed by an experienced information specialist and librarian (MG) in collaboration with the authors. The database search has been carried out by MG who is experienced in the literature search for systematic reviews. The individual search strings for all databases including the number of hits were included as **Additional file 2**. Further, the searches were registered on searchRxiv https://searchrxiv.org/, and they are accessible via the following links:

- Cochrane Library https://doi.org/10.1079/searchRxiv.2024.00438
- Medline https://doi.org/10.1079/searchRxiv.2024.00440
- CINAHL https://doi.org/10.1079/searchRxiv.2024.00437
- Embase https://doi.org/10.1079/searchRxiv.2024.00439
- PsycInfo: https://doi.org/10.1079/searchRxiv.2024.00441

### Selection of studies and data management

After searching the databases and deduplication the identified references will be uploaded to Covidence. Covidence is a software for managing systematic literature reviews (www.covidence.org) where the study selection will take place. The abstract screening will be done by two members of the research team (VP, VE). The first 50 abstracts will be screened by both parties using Covidence. Afterwards, discrepancies will be revised, and a third party (BL, UK, UH) will be involved if disagreements should occur. Upon satisfactory agreement, the remaining studies will be divided and screened by only one person to keep the workload on a manageable level. Otherwise, we will continue double screening and will have another revision after additional 50 abstracts have been screened. Emerging questions and uncertainties will be discussed with each other and a third party. The full text screening will be done by two members of our research team (VP, VE). They will not be blinded to names and affiliations of the authors and journal titles. Arising questions will be discussed with the research team to reach consensus. In case of a publication containing a narrative or systematic review in which not all included studies coincide with our inclusion criteria, we will check if the compatible studies are part of our dataset. If the full text cannot be found, the corresponding author will be contacted to obtain the full text. If the authors do not reply within one month, the reference will be excluded. If the full text is not available and the contact information regarding the corresponding author cannot be found, the study will be excluded. We will manage multiple reports from the same study by documenting that there are overlapping publications. Data of these reports will be handled as stated in the section “Data extraction”. During full text screening, excluded studies as well as their reason for exclusion will be listed in a document.

### Data extraction

The data from the included studies will be extracted using a piloted data extraction form (data extraction form from Covidence will be exported into a comma-separated value file). The members of our research team with biostatistics/methodological expertise (UH, LB, SS) will pilot these forms on five studies and adapt if necessary. The proposal of the draft of the data extraction form is added as **Additional file 3**. Regarding the study population we will extract information on the study population, including gestational age, number of twins, total number of children investigated in the study and further demographic information such as birth weight, age of the mother at birth, length of the hospital stay, the age and the corrected age of the children at baseline at time of intervention. We will record the described interventions, and if applicable also information on standard of care or placebo. Further, we will extract primary and secondary outcomes such as Infant Motor Profile (IMP), Alberta Infant Motor Scale (AIMS), Bayley Scales of Infant and Toddler Development (BSID) etc. For more detailed information on the data extraction we refer to our proposal of the draft of the data extraction form. The data extraction will be done independently by multiple members of the team (VP, SS, UH, UK, LE, LK). The data extraction will follow guidance from a recent paper (12). Any questions arising will be discussed within the research team until consensus is reached. If we have multiple reports from the same study with similar outcomes, we will use the report that corresponds best to 12 months follow-up duration for meta-analysis. If there are two studies with the same time interval as compared to the 12-months follow-up duration (e.g. follow up at 9 and at 15 months), we will incorporate the later one.

For data extraction, publications must report on a minimum set of information in order to be eligible for meta-analysis. The minimum set of information includes a definition of a follow-up timeframe, and the outcome must have been operationalized. Therefore, narrative summaries of findings, or qualitative interviews would not be considered for data extraction and meta-analysis.

### Dealing with missing data

If relevant data is missing in the full text article, the corresponding authors will be contacted to obtain this information. If the authors do not reply within one month, we will use the published data and note that relevant data is missing.

### Assessment of methodological quality and risk of bias (RoB) of included studies

The quality assessment and RoB assessment will be performed by the member of the research team who extracted the data of the respective article (VP, SS, UH, UK, LE and LK). A second party will be involved if questions arise. It will be done at study level.

For observational studies and case series the appropriate SIGN checklist (https://www.sign.ac.uk/what-we-do/methodology/checklists/) will be adapted by members of the research team with biostatistics / methodological expertise (UH, LB, SS) and will be used to perform the RoB assessment. The studies will be coded as “high quality”, “acceptable” or “low quality”. The quality of the study (apart from “unacceptable/reject”) will not be used as weight in the meta-analysis but it will be taken into consideration when discussing the results.

For randomized controlled trials Version 2 of the Cochrane risk-of-bias tool for randomized trials (RoB2) (https://methods.cochrane.org/bias/resources/rob-2-revised-cochrane-risk-bias-tool-randomized-trials) will be used to perform the RoB assessment. The RoB will be coded as “low”, “some concerns” or “high”. The proposal of the draft of the RoB assessment form is added as **Additional file 4**.

### Strategy for data synthesis

If available, we will meta-analyze the results of studies reporting on the same or comparable interventions to standard of care or no structured transition to home interventions. By comparable or similar interventions, we refer to categories such as, e.g., breastfeeding, kangaroo care, NIDCAP (Neonatal Individualised Developmental Care Assessment Program), music therapy, e-health devices, telemedicine, motor development stimulation or family centered care interventions.

Depending on the outcome distribution, different effect measures will be calculated for the individual study and pooled. This includes mean differences or standardized mean differences for continuous outcomes on the same scale or on different scales, as well as risk ratios or risk differences for binary outcomes. We will also summarize studies which report on pre- and post-implementation of specific interventions as well as single group studies.

Random effects meta-analysis models will be used. The heterogeneity variance parameter τ2 will be estimated using restricted maximum likelihood (REML), and DerSimonian-Laird (DL) as sensitivity analysis. Results of the studies will be pooled if three or more studies report on the same outcome. If less studies report on the same outcome, we will report a narrative synthesis. The Hartung-Knapp-Sidik-Jonkman method will be applied in order to adjust the standard errors of the estimated coefficients to account for the uncertainty in the estimate of the amount of residual heterogeneity (13). If the number of studies in a meta-analysis is small, Bayesian methods may be used in an additional sensitivity analysis.

Prediction intervals will be calculated. We do not plan to exclude studies because of high risk of bias, but risk of bias will be evaluated in a meta-regression if the number of studies is large enough. The R programming language will be used for data analysis (14), in combination with the metafor package for meta-analysis. All analyses will be done in a fully scripted way and using dynamic reporting for reproducibility.

### Effect measures

For single arm studies the summary estimates of proportions will be reported for binary outcomes and the summary estimates of the mean will be reported for continuous outcomes. For randomized controlled trials (RCTs), or cohort studies with a comparator group we will use risk ratios or odds ratios for binary outcomes, rate ratio for outcomes with a Poisson distribution, and standardized mean differences or mean differences for continuous outcomes as effect measures.

### Subgroup analyses

If there is enough information, we plan to form and analyze the following subgroups: parental sex, sex of the infant, different durations of follow-up, duration of hospital stay before the transition to home, adjusted age of the infant when transitioning to home. If possible, we will perform a meta-regression for the duration of the hospital stay and for the parental sex.

### Assessment of meta-bias

Funnel plots will be used for the evaluation of publication/reporting bias. Meta-regression will be applied if the number of studies is large enough in order to quantify potential sources of meta-bias.

### Confidence in cumulative evidence

The GRADE framework (https://bestpractice.bmj.com/info/us/toolkit/learn-ebm/what-is-grade/) will be used to present the summary of evidence, in those instances where a pooled summary effect can be calculated for identical or similar interventions as compared to standard-of-care.

### Amendments

If we have to make amendments to the protocol, the date, the reasoning and an exact description of the change will be provided in the final report of the systematic review.

## Discussion

The results of this systematic review will aid and support clinical decision making by providing an overview of existing interventions for the transition from hospital to home process for very preterm-born infants and their parents.

The initial screening has revealed significant heterogeneity across studies concerning the types of interventions. The different interventions can be categorized into subgroups according to their focus areas, including for example, breastfeeding, kangaroo care, NIDCAP (Neonatal Individualised Developmental Care Assessment Program), music therapy, e-health devices, telemedicine, motor development stimulation or family centered care.

We expect the results of the systematic review to provide guidance for the effectiveness of different interventions on relevant outcomes for infants and their parents. The intervention(s) with the most promising effects, and based on the GRADE evaluation of evidence, will then be the basis of a randomized controlled trial which will be designed by members of our research team.

## Supporting information

PRISMA-P checklist

Searchstring documentation

Data extraction proposal

RoB form proposal

## Data Availability

Not applicable - this is a systematic review protocol.

## List of abbreviations

SAVE-T Study: Swiss-Austrian VEry Preterm Infant Transition Study
WHO: World Health Organization
RCT: randomized controlled trial
NIDCAP: Neonatal Individualised Developmental Care Assessment Program
NICU: Neonatal Intensive Care Unit
IMP: Infant Motor Profile
AIMS: Alberta Infant Motor Scale
BSID: Bayley Scales of Infant and Toddler Development
RoB: Risk of bias

## Declarations

### Ethics approval and consent to participate

Not applicable.

### Consent for publication

All authors have agreed to submission and publication of the protocol.

### Availability of data and materials

Not applicable. All data and materials upon completion of the systematic review will be made available on a suitable website (OSF or Zenodo).

### Authors’ contribution

VP and VE drafted the protocol. All authors contributed to the finalization of the protocol. The search strategy was developed by MG in collaboration with the authors. BL and UK provided expertise regarding the eligibility criteria and medical background. UH, LB and SS drafted the meta-analysis methods section and provided methodological expertise. All authors have agreed on the final version. BL is the guarantor of the study.

## Acknowledgements

none

## Author information

Child Development Center, Children’s Research Center, University Children’s Hospital, Switzerland

Pediatric Department, Geneva University Hospital, Switzerland

Department of Biostatistics, at Epidemiology, Biostatistics and Prevention Institute (EBPI), University of Zurich, Switzerland

Medical University of Innsbruck, Austria

Department of Pediatrics II (Neonatology), Medical University of Innsbruck, Austria

University Library, University of Zurich, Switzerland

