## Supplementary material for "Swiss-Austrian VEry Preterm Infant Transition Systematic Review (SAVE-T): Transition to home: improving outcomes of children and families - a systematic review protocol": PRISMA-P checklist

**PRISMA-P (Preferred Reporting Items for Systematic review and Meta-Analysis Protocols) 2015 checklist: recommended items to address in a systematic review protocol***

| Section and topic | Item No | Checklist item |
| --- | --- | --- |
| **ADMINISTRATIVE INFORMATION** | | |
| Title: | | |
| Identification | 1a | Identify the report as a protocol of a systematic review<br>➔ see title |
| Update | 1b | If the protocol is for an update of a previous systematic review, identify as such<br>➔ not applicable |
| Registration | 2 | If registered, provide the name of the registry (such as PROSPERO) and registration number<br>➔ see abstract; systematic review registration and methods/design |
| Authors: | | |
| Contact | 3a | Provide name, institutional affiliation, e-mail address of all protocol authors; provide physical mailing address of corresponding author<br>➔ see title page |
| Contributions | 3b | Describe contributions of protocol authors and identify the guarantor of the review<br>➔ see declarations; authors' contribution |
| Amendments | 4 | If the protocol represents an amendment of a previously completed or published protocol, identify as such and list changes; otherwise, state plan for documenting important protocol amendments<br>➔ see methods/design; amendments |
| Support: | | |
| Sources | 5a | Indicate sources of financial or other support for the review<br>➔ not applicable |
| Sponsor | 5b | Provide name for the review funder and/or sponsor<br>➔ not applicable |
| Role of sponsor or funder | 5c | Describe roles of funder(s), sponsor(s), and/or institution(s), if any, in developing the protocol<br>➔ not applicable |
| **INTRODUCTION** | | |
| Rationale | 6 | Describe the rationale for the review in the context of what is already known<br>➔ see background |
| Objectives | 7 | Provide an explicit statement of the question(s) the review will address with reference to participants, interventions, comparators, and outcomes (PICO) |

| | | |
|---|---|---|
| | | ➔ see objectives |
| **METHODS** | | |
| Eligibility criteria | 8 | Specify the study characteristics (such as PICO, study design, setting, time frame) and report characteristics (such as years considered, language, publication status) to be used as criteria for eligibility for the review<br>➔ methods/design; eligibility criteria, study design, year of publication, language, participants & types of interventions and exposures |
| Information sources | 9 | Describe all intended information sources (such as electronic databases, contact with study authors, trial registers or other grey literature sources) with planned dates of coverage<br>➔ see methods/design; electronic database search |
| Search strategy | 10 | Present draft of search strategy to be used for at least one electronic database, including planned limits, such that it could be repeated<br>➔ see methods/design; electronic database search & search strategy |
| Study records: | | |
| Data management | 11a | Describe the mechanism(s) that will be used to manage records and data throughout the review<br>➔ see methods/design; selection of studies and data management |
| Selection process | 11b | State the process that will be used for selecting studies (such as two independent reviewers) through each phase of the review (that is, screening, eligibility and inclusion in meta-analysis)<br>➔ see methods/design; selection of studies and data management |
| Data collection process | 11c | Describe planned method of extracting data from reports (such as piloting forms, done independently, in duplicate), any processes for obtaining and confirming data from investigators<br>➔ see methos/design; data extraction |
| Data items | 12 | List and define all variables for which data will be sought (such as PICO items, funding sources), any pre-planned data assumptions and simplifications<br>➔ see methods/design; data extraction |
| Outcomes and prioritization | 13 | List and define all outcomes for which data will be sought, including prioritization of main and additional outcomes, with rationale<br>➔ see methods/design; primary outcomes & secondary outcomes |
| Risk of bias in individual studies | 14 | Describe anticipated methods for assessing risk of bias of individual studies, including whether this will be done at the outcome or study level, or both; state how this information will be used in data synthesis<br>➔ see methods/design; assessment of methodological quality and risk of bias of included studies |
| Data synthesis | 15a | Describe criteria under which study data will be quantitatively synthesised<br>➔ see methods/design; strategy for data synthesis |
| | 15b | If data are appropriate for quantitative synthesis, describe planned summary measures, methods of handling data and methods of combining data from studies, including any planned exploration of consistency (such as $I^2$, Kendall's $\tau$)<br>➔ see methods/design; strategy for data synthesis |

| | | |
|---|---|---|
| | 15c | Describe any proposed additional analyses (such as sensitivity or subgroup analyses, meta-regression)<br>➔ see methods/design; subgroup analyses |
| | 15d | If quantitative synthesis is not appropriate, describe the type of summary planned<br>➔ see methods/design; strategy for data synthesis |
| Meta-bias(es) | 16 | Specify any planned assessment of meta-bias(es) (such as publication bias across studies, selective reporting within studies)<br>➔ see methods/design; assessment of meta-bias |
| Confidence in cumulative evidence | 17 | Describe how the strength of the body of evidence will be assessed (such as GRADE)<br>➔ see methods/design; confidence in cumulative evidence |

*** It is strongly recommended that this checklist be read in conjunction with the PRISMA-P Explanation and Elaboration (cite when available) for important clarification on the items. Amendments to a review protocol should be tracked and dated.** **The copyright for PRISMA-P (including checklist) is held by the PRISMA-P Group and is distributed under a Creative Commons Attribution Licence 4.0.**

*From: Shamseer L, Moher D, Clarke M, Ghersi D, Liberati A, Petticrew M, Shekelle P, Stewart L, PRISMA-P Group. Preferred reporting items for systematic review and meta-analysis protocols (PRISMA-P) 2015: elaboration and explanation. BMJ. 2015 Jan 2;349(jan02 1):g7647.*
