## Supplementary material for "Swiss-Austrian VEry Preterm Infant Transition Systematic Review (SAVE-T): Transition to home: improving outcomes of children and families - a systematic review protocol": Data extraction proposal

PROPOSAL OF THE DRAFT OF THE DATA EXTRACTION FORM

### Study Demographics

**reviewer**

Your initials,
e.g. XY

**title**

Title of study that the data are extracted from

**author**

Surname of lead/first author

**publication_year**

Year of publication

**journal**

Name of the journal, the study has been published in.
Do NOT use abbreviations.

**region**

Region in which the study was mainly conducted (the patient were from)

1. Africa
2. Australia / New Zealand
3. Europe
4. Central & South America
5. North America (USA, Canada)
6. East Asia (Chian, Taiwan, Hongkong, Japan, South Korea, North Korea)
7. South East Asia (Brunei, Cambodia, East Timor, Indonesia, Laos, Malaysia, Myanmar, Philippines, Singapore, Thailand, Vietnam)
8. Rest of Asia (e.g. India, Kirgistan)
9. Middle East (Egypt, Iran, Turkey, Saudi Arabia, Yemen, Syria, UAE, Israel, Jordan, Palestine, Lebanon, Oman, Kuwait, Qatar, Bahrain, Cyprus)
10. Not reported
11. Other

**study_design**

Design of the study

1. Randomised controlled trial
2. Observational cohort study: Prospective
3. Observational cohort study: Retrospective
4. Case series with more than 10 cases
5. Case-control study
6. Systematic review
7. Qualitative research
8. Other

**date_start**

Start date of the recruitment.

Format: mm.yyyy
e.g. 02.2005

**date_end**

End date of the recruitment in the study.

Format: mm.yyyy
e.g. 12.2007

**multicenter**

Is the study conducted across multiple centers?

1. Yes
2. No

### Study Population

#### Participants

**GA_min**

Minimum of the GESTATIONAL AGE of infants in the study.

Format: weeks, days
e.g. 28, 0 or 32, 6

If only weeks are specified, use "x" as a placeholder for the days
e.g. 28, x

**GA_max**

Maximum of the GESTATIONAL AGE of infants in the study.

Format: weeks, days
e.g. 28, 0 or 32, 6

If only weeks are specified, use "x" as a placeholder for the days
e.g. 28, x

**n_total**

Total number of children investigated in the study

Use the total sample size: The sample size that does not account for any grouping or stratification.

**demographics_baseline**

Fill in all given patient characteristics in the following table.

If the characteristics are given per intervention group, than add what is possible in this table and add all further information in later tables, that go down onto the level of the groups.

Only enter pure numbers and no units. The units for the different characteristics are:
- birth weight (grams)
- gestational age (weeks): Add days as decimals, e.g. 30 weeks and 1 day = 30.14)
- age of mother at birth (years)
- length of stay in hospital (days)
- age of children at baseline of intervention (weeks)
- corrected age of children at baseline of intervention (weeks)

If the minimal and maximal value of a variable are given, fill them into the placeholders for range_min, range_max.

| | mean | SD | median | IQR_lower | IQR_upper | range_min | range_max |
|---|---|---|---|---|---|---|---|
| **birth_weight** | | | | | | | |
| **gestational_age** | | | | | | | |
| **age_mother_birth** | | | | | | | |
| **length_of_stay** | | | | | | | |
| **age_children_int** | | | | | | | |
| **corr_age_children_int** | | | | | | | |

### Study Groups

**groups**

How many different study groups are there in the study? E.g. control arm and different intervention arms. Please note that the control arm also counts. If the study only includes one treatment or is a qualitative analysis of one intervention please choose "1" as answer.

1. 1
2. 2
3. 3
4. 4
5. Other

### Group 1: define group 1

**group1_definition**

Select all appropriate interventions for GROUP 1.

1. No Intervention
2. Control
3. Sham Therapy
4. Standard of Care
5. Audio / Language
6. Booklet Device
7. Breast Feeding
8. eHealth
9. Face-to-face
10. Kangaroo Mother Care

11. Music Theraphy
12. Nidcap
13. Nutrition
14. Parental Advice/ Education
15. Supporting Play Exploration and Early Development Intervention (SPEEDI)
16. Telemedicine
17. Other

**group1_explanation**

Add any further information about the intervention in GROUP 1,
e.g. daily music therapy conducted at home.

**demographics_group1**

Fill in all patient characteristics for GROUP 1 given in the following table.

Only enter pure numbers and no units. The units for the different characteristics are:
- birth weight (grams)
- gestational age (weeks): Add days as decimals, e.g. 30 weeks and 1 day = 30.14)
- age of mother at birth (years)
- length of stay in hospital (days)
- age of children at baseline of intervention (weeks)
- corrected age of children at baseline of intervention (weeks)

If the minimal and maximal value of a variable are given, fill them into the placeholders for range_min, range_max.

| | mean | SD | median | IQR_lower | IQR_upper | range_min | range_max |
|---|---|---|---|---|---|---|---|
| **birth_weight** | | | | | | | |
| **gestational_age** | | | | | | | |
| **age_mother** | | | | | | | |
| **length_of_stay** | | | | | | | |
| **age_children_int** | | | | | | | |
| **corr_age_children_int** | | | | | | | |

### Group 2: define group 2

**group2_definition**

Select all appropriate interventions for GROUP 2.

1. No Intervention
2. Control
3. Sham Therapy
4. Standard of Care
5. Audio / Language
6. Booklet Device
7. Breast Feeding
8. eHealth

9. Face-to-face
10. Kangoroo Mother Care
11. Music Therapy
12. Nidcap
13. Nutrition
14. Parental Advice/ Education
15. Supporting Play Exploration and Early Development Intervention (SPEEDI)
16. Telemedicine
17. Other

**group2_explanation**

Add any further information about intervention in GROUP 2,
e.g. daily music therapy conducted at home.

**group2_demographics**

Fill in all patient characteristics for GROUP 2 given in the following table.

Only enter pure numbers and no units. The units for the different characteristics are:
- birth weight (grams)
- gestational age (weeks): Add days as decimals, e.g. 30 weeks and 1 day = 30.14)
- age of mother at birth (years)
- length of stay in hospital (days)
- age of children at baseline of intervention (weeks)
- corrected age of children at baseline of intervention (weeks)
If the minimal and maximal value of a variable are given, fill them into the placeholders for range_min, range_max.

| | mean | SD | median | IQR_lower | IQR_upper | range_min | range_max |
|---|---|---|---|---|---|---|---|
| **birth_weight** | | | | | | | |
| **gestational_age** | | | | | | | |
| **age_mother** | | | | | | | |
| **length_of_stay** | | | | | | | |
| **age_children_int** | | | | | | | |
| **corr_age_children_int** | | | | | | | |

### Group 3: define group 3

**group3_defintion**

Select all appropriate interventions for GROUP 3.

1. No Intervention
2. Control
3. Sham Therapy
4. Standard of Care
5. Audio / Language
6. Booklet Device
7. Breast Feeding

8. eHealth
9. Face-to-face
10. Kangoroo Mother Care
11. Music Therapy
12. Nidcap
13. Nutrition
14. Parental Advice/ Education
15. Supporting Play Exploration and Early Development Intervention (SPEEDI)
16. Telemedicine
17. Other

**group3_explanation**

Add any further information about the intervention in GROUP 3,
e.g. daily music therapy conducted at home.

**group3_demographics**

Fill in all patient characteristics for the GROUP 3 given in the following table.

Only enter pure numbers and no units. The units for the different characteristics are:
- birth weight (grams)
- gestational age (weeks): Add days as decimals, e.g. 30 weeks and 1 day = 30.14)
- age of mother at birth (years)
- length of stay in hospital (days)
- age of children at baseline of intervention (weeks)
- corrected age of children at baseline of intervention (weeks)

If the minimal and maximal value of a variable are given, fill them into the placeholders for range_min, range_max.

| | mean | SD | median | IQR_lower | IQR_upper | range_min | range_max |
|---|---|---|---|---|---|---|---|
| **birth_weight** | | | | | | | |
| **gestational_age** | | | | | | | |
| **age_mother** | | | | | | | |
| **length_of_stay** | | | | | | | |
| **age_children_int** | | | | | | | |
| **corr_age_children_int** | | | | | | | |

### group 4: define group 4

**group4_definition**

Select all appropriate interventions for GROUP 4.

1. No Intervention
2. Control
3. Sham Therapy
4. Standard of Care
5. Audio / Language

6. Booklet Device
7. Breast Feeding
8. eHealth
9. Face-to-face
10. Kangaroo Mother Care
11. Music Theraphy
12. Nidcap
13. Nutrition
14. Parental Advice/ Education
15. Supporting Play Exploration and Early Development Intervention (SPEEDI)
16. Telemedicine
17. Other

**group4_explanation**

Add any further information about the intervention in GROUP 4,
e.g. daily music therapy conducted at home.

**group4_demographics**

Fill in all patient characteristics for GROUP 4 given in the following table.

Only enter pure numbers and no units. The units for the different characteristics are:
- birth weight (grams)
- gestational age (weeks): Add days as decimals, e.g. 30 weeks and 1 day = 30.14)
- age of mother at birth (years)
- length of stay in hospital (days)
- age of children at baseline of intervention (weeks)
- corrected age of children at baseline of intervention (weeks)

If the minimal and maximal value of a variable are given, fill them into the placeholders for range_min, range_max.

| | mean | SD | median | IQR_lower | IQR_upper | range_min | range_max |
|---|---|---|---|---|---|---|---|
| **birth_weight** | | | | | | | |
| **gestational_age** | | | | | | | |
| **age_mother** | | | | | | | |
| **length_of_stay** | | | | | | | |
| **age_children_int** | | | | | | | |
| **corr_age_children_int** | | | | | | | |

### Baseline parameters for all study arms

**n_groups**

Fill in the sample size per GROUP at the beginning of the study (i.e., at baseline).

If available take the sample size from the table with the baseline characteristics (Table 1).

| | n |
|---|---|
| **group1** | |
| **group2** | |
| **group3** | |
| **group4** | |

**n_sex**

Fill in the sample size per SEX and GROUP at the beginning of the study (i.e., at time of randomization, at baseline)

| | female | male |
|---|---|---|
| **n** | | |
| **group1** | | |
| **group2** | | |
| **group3** | | |
| **group4** | | |

**twins**

NUMBER OF TWINS at the beginning of the study for each group.

Fill in the total amount of twins and not the number of twin couples,
e.g., if there are totally 5 twin couples, n would be 10 as every twin counts.

| | n |
|---|---|
| **overall** | |
| **group1** | |
| **group2** | |
| **group3** | |
| **group4** | |

**complications**

Fill in the number of children with complications at the beginning of the study for each group.

| | n |
|---|---|
| **overall** | |
| **group1** | |
| **group2** | |
| **group3** | |

| | n |
|---|---|
| **group4** | |

**complications_explanation**

What is/are the complications?

Format: comma-separated
e.g., xx, yy, zz

### Outcomes

**primary_outcome**

Select the primary outcome.

1. Infant Motor Profile (IMP)
2. Alberta Infant Motor Scale (AIMS)
3. Teller Acuity Cards
4. Griffiths
5. Bayley Scales of Infant and Toddler Development (BSID)
6. Edinburgh Edinburgh Postnatal Depression Scale
7. Infant Behavioral Assessment (IBA)
8. Parenting Stress Index short form (PSI-SF)
9. Neurobehavioural Assessment of the Preterm Infant (NAPI)
10. Nursing Child Assessment Teaching Scale (NCATS)
11. Home Observation for Measurement of the Environment (HOME)
12. Behavioral Rating Scale (BRS)
13. Other

**primary_outcome_scale_lower**

If the primary outcome is a scale/score, indicate the LOWER limit of the scale/score (numeric).

**primary_outcome_scale_upper**

If the primary outcome is a scale/score, please indicate the UPPER limit of the scale/score (numeric).

**primary_outcome_direction**

In which direction does the outcome (e.g. score) go?

1. score: higher value = better condition of child
2. score: higher value = worse condition of child
3. binary outcome: event = positive for child
4. binary outvome: event = negative for child

**primary_outcome_effect_measure_single_group**

Select the effect measure of the primary outcome, if ONE group is given.

1. Mean
2. Change Score Mean
3. Proportion

**primary_outcome_effect_measure**

Select the effect measure of the primary outcome, if MULTIPLE groups are given.

1. Mean
2. Mean Difference
3. Change Score Mean
4. Mean Difference in Change Score
5. Proportion
6. Risk Difference
7. Odds Ratio
8. Risk Ratio
9. Hazard Ratio
10. only p-values or test statistics
11. Other

**secondary_outcome**

Select the main secondary outcome.

1. Infant Motor Profile (IMP)
2. Alberta Infant Motor Scale (AIMS)
3. Teller Acuity Cards
4. Griffiths
5. Bayley Scales of Infant and Toddler Development (BSID)
6. Edinburgh Postnatal Depression Scale
7. Infant Behavioral Assessment (IBA)
8. Parenting Stress Index short form (PSI-SF)
9. Neurobehavioural Assessment of the Preterm Infant (NAPI)
10. Nursing Child Assessment Teaching Scale (NCATS)
11. Home Observation for Measurement of the Environment (HOME)
12. Behavioral Rating Scale (BRS)
13. Other

**secondary_outcome_scale_lower**

If the secondary outcome is a scale/score, please indicate the LOWER limit of the scale/score (numeric).

**secondary_outcome_scale_upper**

If the secondary outcome is a scale/score, please indicate the UPPER limit of the scale/score (numeric).

**secondary_outcome_direction**

In which direction does the outcome (e.g. score) go?

1. score: higher value = better condition of child
2. score: higher value = worse condition of child
3. binary outcome: event = positive for child
4. binary outcome: event = negative for child

**secondary_outcome_effect_measure_single_group**

Select the effect measure of the primary outcome, if ONE group is given.

1. Mean
2. Change Score Mean
3. Proportion

**secondary_outcome_effect_measure**

Select the effect measure of the primary outcome, if MULTIPLE groups are given.

1. Mean
2. Mean Difference
3. Change Score Mean
4. Mean Difference in Change Score
5. Proportion
6. Risk Difference
7. Odds Ratio
8. Risk Ratio
9. Hazard Ratio
10. only p-values or test statistics
11. Other

**secondary_outcome_further**

Enter all further secondary outcomes.

Format: comma-separated
e.g., xx, yy, zz

**followup_timepoint**

Add the follow-up time points in months.

Format: months comma-separated,
e.g.: 0.5, 1, 2, 4, 8, 12, 24

**main_followup_timepoint**

Add the follow-up time point (format: months) which is the closest to 12 months.
This will be our main follow-up time point (see PROSPERO).

Format: time point in months,
e.g. 8 or 14.

When two follow-ups have the same distance to the 12 months, than always choose the second one. E.g. 10 and 14 month follow-up given, chose the 14 month follow-up as main follow-up.

### Results: Primary Outcome

#### Binary Outcomes: Fill in only if primary outcome is binary

**primary_outcome_fu_binary**

Fill in the results of the PRIMARY OUTCOME per GROUP at the main follow-up defined before.

Add the sample size per group at the given follow-up.

| | n | n_Event | Proportion |
|---|---|---|---|
| **group1** | | | |
| **group2** | | | |
| **group3** | | | |
| **group4** | | | |

**primary_outcome_effect_binary_unadjusted**

Fill in the UNADJUSTED ESTIMATES and the corresponding confidence intervals (95%) and p-values.

NOTE: Unadjusted means the raw effect (most often the case if nothing is written).

| | estimate | CI_lower | CI_upper | p_value |
|---|---|---|---|---|
| **group1_vs_refgroup2** | | | | |
| **group1_vs_refgroup3** | | | | |
| **group1_vs_refgroup4** | | | | |
| **group2_vs_refgroup1** | | | | |
| **group2_vs_refgroup3** | | | | |
| **group2_vs_refgroup4** | | | | |
| **group3_vs_refgroup1** | | | | |
| **group3_vs_refgroup2** | | | | |
| **group3_vs_refgroup4** | | | | |
| **group4_vs_refgroup1** | | | | |
| **group4_vs_refgroup2** | | | | |
| **group4_vs_refgroup3** | | | | |

**primary_outcome_effect_binary_adjusted**

Fill in the ADJUSTED ESTIMATES and the corresponding confidence intervals (95%) and p-values.

NOTE: Adjusted is most often indicated with a * or something similar. E.g. *adjusted for age.

| | estimate | CI_lower | CI_upper | p_value |
|---|---|---|---|---|
| **group1_vs_refgroup2** | | | | |

| | estimate | CI_lower | CI_upper | p_value |
|---|---|---|---|---|
| **group1_vs_refgroup3** | | | | |
| **group1_vs_refgroup4** | | | | |
| **group2_vs_refgroup1** | | | | |
| **group2_vs_refgroup3** | | | | |
| **group2_vs_refgroup4** | | | | |
| **group3_vs_refgroup1** | | | | |
| **group3_vs_refgroup2** | | | | |
| **group3_vs_refgroup4** | | | | |
| **group4_vs_refgroup1** | | | | |
| **group4_vs_refgroup2** | | | | |
| **group4_vs_refgroup3** | | | | |

**primary_outcome_effect_binary_adjusted_variable**

What has been adjusted for?

Format: comma separated,
e.g. gestational age, sex, maternal education,...

**Continous Outcome: Fill in only if primary outcome is continous**

**primary_outcome_baseline_continuous**

Fill in the results for the PRIMARY OUTCOME per GROUP at baseline.

Add the sample size per group at baseline.

| | n | mean | SE | SD | CI_lower | CI_upper | median | IQR_lower | IQR_upper |
|---|---|---|---|---|---|---|---|---|---|
| **group1** | | | | | | | | | |
| **group2** | | | | | | | | | |
| **group3** | | | | | | | | | |
| **group4** | | | | | | | | | |

**primary_outcome_followup_continuous**

Fill in the results for the PRIMARY OUTCOME per GROUP at the main follow-up defined before.

Add the sample size per group at given follow-up.

| | n | mean | SE | SD | CI_lower | CI_upper | median | IQR_lower | IQR_upper |
|---|---|---|---|---|---|---|---|---|---|
| **group1** | | | | | | | | | |
| **group2** | | | | | | | | | |
| **group3** | | | | | | | | | |
| **group4** | | | | | | | | | |

**primary_outcome_effect_continuous_unadjusted**

Fill in the UNADJUSTED ESTIMATES and the corresponding confidence intervals (95%) and p-values.

NOTE: Unadjusted means the raw effect (most often the case if nothing is written).

| | Estimate | CI_lower | CI_upper | p_value |
|---|---|---|---|---|
| **group1_vs_refgroup2** | | | | |
| **group1_vs_refgroup3** | | | | |
| **group1_vs_refgroup4** | | | | |
| **group2_vs_refgroup1** | | | | |
| **group2_vs_refgroup3** | | | | |
| **group2_vs_refgroup4** | | | | |
| **group3_vs_refgroup1** | | | | |
| **group3_vs_refgroup2** | | | | |
| **group3_vs_refgroup4** | | | | |
| **group4_vs_refgroup1** | | | | |
| **group4_vs_refgroup2** | | | | |
| **group4_vs_refgroup3** | | | | |

**primary_outcome_effect_continuous_adjusted**

Fill in the ADJUSTED ESTIMATES and the corresponding confidence intervals (95%) and p-values.

NOTE: Adjusted is most often indicated with a * or something similar. E.g. *adjusted for age.

| | Estimate | CI_lower | CI_upper | p_value |
|---|---|---|---|---|
| **group1_vs_refgroup2** | | | | |
| **group1_vs_refgroup3** | | | | |
| **group1_vs_refgroup4** | | | | |
| **group2_vs_refgroup1** | | | | |

| | Estimate | CI_lower | CI_upper | p_value |
|---|---|---|---|---|
| **group2_vs_refgroup3** | | | | |
| **group2_vs_refgroup4** | | | | |
| **group3_vs_refgroup1** | | | | |
| **group3_vs_refgroup2** | | | | |
| **group3_vs_refgroup4** | | | | |
| **group4_vs_refgroup1** | | | | |
| **group4_vs_refgroup2** | | | | |
| **group4_vs_refgroup3** | | | | |

**primary_outcome_effect_continous_adjusted_variable**

What has been adjusted for?

Format: comma separated,
e.g. gestational age, sex, maternal education,...

### Results: Secondary Outcome

#### Binary Outcomes: Fill in only if secondary outcome is binary

**secondary_outcome_followup_binary**

Fill in the results for the SECONDARY OUTCOME per GROUP at the main follow-up defined before.

Add the sample size per group at the given follow-up.

| | n | n_Event | Proportion |
|---|---|---|---|
| **group1** | | | |
| **group2** | | | |
| **group3** | | | |
| **group4** | | | |

**secondary_outcome_effect_binary_unadjusted**

Fill in the UNADJUSTED ESTIMATES and the corresponding confidence intervals (95%) and p-values.

NOTE: Unadjusted means the raw effect (most often the case if nothing is written).

| | Estimate | CI_lower | CI_upper | p_value |
|---|---|---|---|---|
| **group1_vs_refgroup2** | | | | |

| | Estimate | CI_lower | CI_upper | p_value |
|---|---|---|---|---|
| **group1_vs_refgroup3** | | | | |
| **group1_vs_refgroup4** | | | | |
| **group2_vs_refgroup1** | | | | |
| **group2_vs_refgroup3** | | | | |
| **group2_vs_refgroup4** | | | | |
| **group3_vs_refgroup1** | | | | |
| **group3_vs_refgroup2** | | | | |
| **group3_vs_refgroup4** | | | | |
| **group4_vs_refgroup1** | | | | |
| **group4_vs_refgroup2** | | | | |
| **group4_vs_refgroup3** | | | | |

**secondary_outcome_effect_binary_adjusted**

Fill in the ADJUSTED ESTIMATES and the corresponding confidence intervals (95%) and p-values.

NOTE: Adjusted is most often indicated with a * or something similar. E.g. *adjusted for age.

| | Estimate | CI_lower | CI_upper | p_value |
|---|---|---|---|---|
| **group1_vs_refgroup2** | | | | |
| **group1_vs_refgroup3** | | | | |
| **group1_vs_refgroup4** | | | | |
| **group2_vs_refgroup1** | | | | |
| **group2_vs_refgroup2** | | | | |
| **group2_vs_refgroup4** | | | | |
| **group3_vs_refgroup1** | | | | |
| **group3_vs_refgroup2** | | | | |
| **group3_vs_refgroup4** | | | | |
| **group4_vs_refgroup1** | | | | |
| **group4_vs_refgroup2** | | | | |
| **group4_vs_refgroup3** | | | | |

**secondary_outcome_effect_binary_adjusted_variable**

What has been adjusted for?

Format: comma separated,
e.g. gestational age, sex, maternal education, ...

**Continous Outcome: Fill in only if secondary outcome is continous**

**secondary_outcome_baseline_continous**

Fill in the results for the PRIMARY OUTCOME per GROUP at baseline.

Add the sample size per group at baseline.

| | n | Mean | SE | SD | CI_lower | CI_upper | Median | IQR_lower | IQR_upper |
|---|---|---|---|---|---|---|---|---|---|
| **group1** | | | | | | | | | |
| **group2** | | | | | | | | | |
| **group3** | | | | | | | | | |
| **group4** | | | | | | | | | |

**secondary_outcome_followup_continous**

Fill in the results for the PRIMARY OUTCOME per GROUP at the main follow-up defined before.

Add the sample size per group at given follow-up.

| | n | Mean | SE | SD | CI_lower | CI_upper | Median | IQR_lower | IQR_upper |
|---|---|---|---|---|---|---|---|---|---|
| **group1** | | | | | | | | | |
| **group2** | | | | | | | | | |
| **group3** | | | | | | | | | |
| **group4** | | | | | | | | | |

**secondary_outcome_effect_continous_unadjusted**

Fill in the UNADJUSTED ESTIMATES and the corresponding confidence intervals (95%) and p-values.

NOTE: Unadjusted means the raw effect (most often the case if nothing is written).

| | Estimate | CI_lower | CI_upper | p_value |
|---|---|---|---|---|
| **group1_vs_refgroup2** | | | | |
| **group1_vs_refgroup3** | | | | |
| **group1_vs_refgroup4** | | | | |
| **group2_vs_refgroup1** | | | | |
| **group2_vs_refgroup3** | | | | |

| | Estimate | CI_lower | CI_upper | p_value |
|---|---|---|---|---|
| **group2_vs_refgroup4** | | | | |
| **group3_vs_refgroup1** | | | | |
| **group3_vs_refgroup2** | | | | |
| **group3_vs_refgroup4** | | | | |
| **group4_vs_refgroup1** | | | | |
| **group4_vs_refgroup2** | | | | |
| **group4_vs_refgroup3** | | | | |

**secondary_outcome_effect_continous_adjusted**

Fill in the ADJUSTED ESTIMATES and the corresponding confidence intervals (95%) and p-values.

NOTE: Adjusted is most often indicated with a * or something similar. E.g. *adjusted for age.

| | Estimate | CI_lower | CI_upper | p_value |
|---|---|---|---|---|
| **group1_vs_refgroup2** | | | | |
| **group1_vs_refgroup3** | | | | |
| **group1_vs_refgroup4** | | | | |
| **group2_vs_refgroup1** | | | | |
| **group2_vs_refgroup3** | | | | |
| **group2_vs_refgroup4** | | | | |
| **group3_vs_refgroup1** | | | | |
| **group3_vs_refgroup2** | | | | |
| **group3_vs_refgroup4** | | | | |
| **group4_vs_refgroup1** | | | | |
| **group4_vs_refgroup2** | | | | |
| **group4_vs_refgroup3** | | | | |

**secondary_outcome_effect_contionus_adjusted_variable**

What has been adjusted for?

Format: comma separated,
e.g. gestational age, sex, maternal education,...

### Subgroups

**subgroups_yn**

Was the (effect of) primary outcome also reported in subgroups?

1. Yes
2. No

**subgroups**

If the (effect of) the primary outcome has been reported in subgroups, please state the subgroups.

Format: comma separated,
e.g. gestational age, sex, maternal education,...
