## Supplementary material for "Swiss-Austrian VEry Preterm Infant Transition Systematic Review (SAVE-T): Transition to home: improving outcomes of children and families - a systematic review protocol": RoB form proposal

PROPOSAL OF THE DRAFT OF THE RISK OF BIAS ASSESSMENT FORM

### RCT: 1.1 Randomization process

Was the allocation sequence generated randomly?

1. Yes
2. Partially yes
3. Partially no
4. No
5. Can't say
6. Does not apply

*Extractors will also be able to add supporting text to justify their judgements*

### RCT: 1.2 Randomization process

Was the allocation sequence concealed until participants were enrolled and assigned to the groups?

1. Yes
2. Partially yes
3. Partially no
4. No
5. Can't say
6. Does not apply

*Extractors will also be able to add supporting text to justify their judgements*

### RCT: 1.3 Randomization process

Did baseline differences between groups suggest a problem with the randomization process?

1. Yes
2. Partially yes
3. Partially no
4. No

5. Can't say
6. Does not apply

*Extractors will also be able to add supporting text to justify their judgements*

### RCT: 1 Risk-of-bias judgement

From the answers above, is there a risk of bias in the study?

1. Low
2. Some concerns
3. High

*Extractors will also be able to add supporting text to justify their judgements*

### RCT: 2.1 Deviations due to assignment to intervention

Were participants aware of their assigned intervention during the trial?

1. Yes
2. Partially yes
3. Partially no
4. No
5. Can't say
6. Does not apply

*Extractors will also be able to add supporting text to justify their judgements*

### RCT: 2.2 Deviations due to assignment to intervention

Were carers and people delivering the interventions aware of participants' assigned intervention during the trial?

1. Yes
2. Partially yes
3. Partially no
4. No
5. Can't say
6. Does not apply

*Extractors will also be able to add supporting text to justify their judgements*

### RCT: 2.3 Deviations due to assignment to intervention (follow-up if 2.1/2.2 are yes or partially yes)

Were there deviations from the intended intervention that arose because of the trial context?

1. Yes
2. Partially yes
3. Partially no
4. No
5. Can't say
6. Does not apply

*Extractors will also be able to add supporting text to justify their judgements*

### RCT: 2.4 Deviations due to assignment to intervention (follow-up if 2.3 is yes or partially yes)

Were these deviations likely to have affected the outcome?

1. Yes
2. Partially yes
3. Partially no
4. No
5. Can't say
6. Does not apply

*Extractors will also be able to add supporting text to justify their judgements*

### RCT: 2.5 Deviations due to assignment to intervention (follow-up if 2.4 is yes or partially yes)

Were these deviations from intended intervention balanced between groups?

1. Yes
2. Partially yes
3. Partially no

4. No
5. Can't say
6. Does not apply

*Extractors will also be able to add supporting text to justify their judgements*

### RCT: 2.6 Deviations due to assignment to intervention

Was an appropriate analysis used to estimate the effect of the intervention(s)?

1. Yes
2. Partially yes
3. Partially no
4. No
5. Can't say
6. Does not apply

*Extractors will also be able to add supporting text to justify their judgements*

### RCT: 2.7 Deviations due to assignment to intervention (follow-up if 2.6 is no or partially no)

Was there potential for a substantial impact (on the result) of the failure to analyze participants in the group to which they were randomized?

1. Yes
2. Partially yes
3. Partially no
4. No
5. Can't say
6. Does not apply

*Extractors will also be able to add supporting text to justify their judgements*

### RCT: 2 Risk-of-bias judgement

From the answers above, is there a risk of bias in the study?

1. Low

2. Some concerns
3. High

*Extractors will also be able to add supporting text to justify their judgements*

### RCT: 3.3 Deviations due to adhering to intervention (follow-up if 3.1/3.2 are yes or partially yes)

Were important non-protocol interventions balanced across groups?

1. Yes
2. Partially yes
3. Partially no
4. No
5. Can't say
6. Does not apply

*Extractors will also be able to add supporting text to justify their judgements*

### RCT: 3.4 Deviations due to adhering to intervention

Were there failures in implementing the intervention that could have affected the outcome?

1. Yes
2. Partially yes
3. Partially no
4. No
5. Can't say
6. Does not apply

*Extractors will also be able to add supporting text to justify their judgements*

### RCT: 3.5 Deviations due to adhering to intervention

Was there non-adherence to the assigned intervention regimen that could have affected participants' outcomes?

1. Yes
2. Partially yes

3. Partially no
4. No
5. Can't say
6. Does not apply

*Extractors will also be able to add supporting text to justify their judgements*

### RCT: 3.6 Deviations due to adhering to intervention (follow-up if 3.3 is no or partially no OR 2.4 is yes or partially yes)

Was an appropriate analysis used to estimate the effect of adhering to the intervention?

1. Yes
2. Partially yes
3. Partially no
4. No
5. Can't say
6. Does not apply

*Extractors will also be able to add supporting text to justify their judgements*

### RCT: 3 Risk-of-bias judgement

From the answers above, is there a risk of bias in the study?

1. Low
2. Some concerns
3. High

*Extractors will also be able to add supporting text to justify their judgements*

### RCT: 4.1 Missing outcome data

Were data for this outcome available for all, or nearly all, participants randomized?

1. Yes
2. Partially yes
3. Partially no
4. No

5. Can't say
6. Does not apply

*Extractors will also be able to add supporting text to justify their judgements*

### RCT: 4.2 Missing outcome data (follow-up if 4.1 is no or partially no)

Is there evidence that the result was not biased by missing outcome data?

1. Yes
2. Partially yes
3. Partially no
4. No
5. Can't say
6. Does not apply

*Extractors will also be able to add supporting text to justify their judgements*

### RCT: 4.3 Missing outcome data (follow-up if 4.2 is no or partially no)

Could missingness in the outcome depend on its true value?

1. Yes
2. Partially yes
3. Partially no
4. No
5. Can't say
6. Does not apply

*Extractors will also be able to add supporting text to justify their judgements*

### RCT: 4.4 Missing outcome data (follow-up if 4.1 is yes or partially yes)

Is it likely that missingness in the outcome depended on its true value?

1. Yes
2. Partially yes
3. Partially no

4. No
5. Can't say
6. Does not apply

*Extractors will also be able to add supporting text to justify their judgements*

### RCT: 4 Risk-of-bias judgement

From the answers above, is there a risk of bias in the study?

1. Low
2. Some concerns
3. High

*Extractors will also be able to add supporting text to justify their judgements*

### RCT: 5.1 Measurement of Outcome

Was the method of measuring the outcome inappropriate?

1. Yes
2. Partially yes
3. Partially no
4. No
5. Can't say
6. Does not apply

*Extractors will also be able to add supporting text to justify their judgements*

### RCT: 5.2 Measurement of Outcome

Could measurement or ascertainment of the outcome have differed between intervention groups?

1. Yes
2. Partially yes
3. Partially no
4. No
5. Can't say

6. Does not apply

*Extractors will also be able to add supporting text to justify their judgements*

### RCT: 5.3 Measurement of Outcome (follow-up if 5.1 and 5.2 are BOTH no/partially no)

Were outcome assessors aware of the intervention received by study participants?

1. Yes
2. Partially yes
3. Partially no
4. No
5. Can't say
6. Does not apply

*Extractors will also be able to add supporting text to justify their judgements*

### RCT: 5.4 Measurement of Outcome (follow-up if 5.3 is yes or partially yes)

Could assessment of the outcome have been influenced by knowledge of intervention received?

1. Yes
2. Partially yes
3. Partially no
4. No
5. Can't say
6. Does not apply

*Extractors will also be able to add supporting text to justify their judgements*

### RCT: 5.5 Measurement of Outcome (follow-up if 5.4 is yes or partially yes)

Is it likely that assessment of the outcome was influenced by knowledge of intervention received?

1. Yes

2. Partially yes
3. Partially no
4. No
5. Can't say
6. Does not apply

*Extractors will also be able to add supporting text to justify their judgements*

### RCT: 5 Risk-of-bias judgement

From the answers above, is there a risk of bias in the study?

1. Low
2. Some concerns
3. High

*Extractors will also be able to add supporting text to justify their judgements*

### RCT: 6.1 Results

Were the data that produced this result analyzed in accordance with a pre-specified analysis plan that was finalized before unblinded outcome data were available for analysis?

1. Yes
2. Partially yes
3. Partially no
4. No
5. Can't say
6. Does not apply

*Extractors will also be able to add supporting text to justify their judgements*

### RCT: 6.2 Results

Is the numerical result being assessed likely to have been selected, on the basis of the results, from multiple eligible outcome measurements (e.g. scales, definitions, time points) within the outcome domain?

1. Yes
2. Partially yes

3. Partially no
4. No
5. Can't say
6. Does not apply

*Extractors will also be able to add supporting text to justify their judgements*

### RCT: 6.3 Results

Is the numerical result being assessed likely to have been selected, on the basis of the results, from multiple eligible analyses of the data?

1. Yes
2. Partially yes
3. Partially no
4. No
5. Can't say
6. Does not apply

*Extractors will also be able to add supporting text to justify their judgements*

### RCT: 6 Risk-of-bias judgement

From the answers above, is there a risk of bias in the study?

1. Low
2. Some concerns
3. High

*Extractors will also be able to add supporting text to justify their judgements*

### RCT: OVERALL Risk-of-bias judgement

From ALL answers above, is there an OVERALL risk of bias in the study?

1. Low
2. Some concerns
3. High

*Extractors will also be able to add supporting text to justify their judgements*

### OBS: 1.1 General

The study addresses an appropriate and clearly focused question.

1. Yes
2. Partially yes
3. Partially no
4. No
5. Can't say
6. Does not apply

*Extractors will also be able to add supporting text to justify their judgements*

### OBS: 2.1 Subjects

The groups being studied are selected from source populations that are comparable in all respects other than the factor under investigation.

1. Yes
2. Partially yes
3. Partially no
4. No
5. Can't say
6. Does not apply

*Extractors will also be able to add supporting text to justify their judgements*

### OBS: 2.2 Subjects

The study indicates how many of the people asked to take part did so, in each of the groups being studied.

1. Yes
2. Partially yes
3. Partially no
4. No
5. Can't say

6. Does not apply

*Extractors will also be able to add supporting text to justify their judgements*

### OBS: 2.3 Subjects

The likelihood that some eligible subjects might have the outcome at the time of enrollment is assessed and taken into account in the analysis.

1. Yes
2. Partially yes
3. Partially no
4. No
5. Can't say
6. Does not apply

*Extractors will also be able to add supporting text to justify their judgements*

### OBS: 2.4 Subjects

Are the results of this study directly applicable to the patient group targeted?

1. Yes
2. Partially yes
3. Partially no
4. No
5. Can't say
6. Does not apply

*Extractors will also be able to add supporting text to justify their judgements*

### OBS: 2 Risk-of-bias judgement

From the answers above, is there a risk of bias in the study?

1. Low
2. Some concerns
3. High

*Extractors will also be able to add supporting text to justify their judgements*

### OBS: 3.1 Outcome

The outcomes are clearly defined.

1. Yes
2. Partially yes
3. Partially no
4. No
5. Can't say
6. Does not apply

*Extractors will also be able to add supporting text to justify their judgements*

### OBS: 3.2 Outcome

The assessment of outcome is made blind to group assignment. If the study is retrospective this may not be applicable.

1. Yes
2. Partially yes
3. Partially no
4. No
5. Can't say
6. Does not apply

*Extractors will also be able to add supporting text to justify their judgements*

### OBS: 3.3 Outcome

Where blinding was not possible, there is some recognition that knowledge of group assignment could have influenced the assessment of outcome

1. Yes
2. Partially yes
3. Partially no
4. No
5. Can't say

6. Does not apply

*Extractors will also be able to add supporting text to justify their judgements*

### OBS: 3.4 Outcome

Evidence from other sources is used to demonstrate that the method of outcome assessment is valid and reliable.

1. Yes
2. Partially yes
3. Partially no
4. No
5. Can't say
6. Does not apply

*Extractors will also be able to add supporting text to justify their judgements*

### OBS: 3 Risk-of-bias judgement

From the answers above, is there a risk of bias in the study?

1. Low
2. Some concerns
3. High

*Extractors will also be able to add supporting text to justify their judgements*

### OBS: 4.1 Confounding and Statistics

The main potential confounders are identified and taken into account in the design and analysis.

1. Yes
2. Partially yes
3. Partially no
4. No
5. Can't say
6. Does not apply

*Extractors will also be able to add supporting text to justify their judgements*

### OBS: 4.2 Confounding and Statistics

Have confidence intervals been provided?

1. Yes
2. Partially yes
3. Partially no
4. No
5. Can't say
6. Does not apply

*Extractors will also be able to add supporting text to justify their judgements*

### OBS: 4 Risk-of-bias judgement

From the answers above, is there a risk of bias in the study?

1. Low
2. Some concerns
3. High

*Extractors will also be able to add supporting text to justify their judgements*

### OBS: OVERALL Risk-of-bias judgement

From all answers above, is there a risk of bias in the study?

1. Low
2. Some concerns
3. High

*Extractors will also be able to add supporting text to justify their judgements*
